# A proteogenomic atlas of idiopathic pulmonary arterial hypertension reveals sex-dimorphic mechanisms and potential novel therapeutic targets

**DOI:** 10.64898/2025.12.11.25341960

**Authors:** Xinjie Lin, YangChang Zhang, Yang Pu, Tao Lei, Kai Ma, Qiyu He, Zheng Dou, Yuze Liu, Dengyuan Liu, Yinge He, Yanshang Wang, Xiaojiao Zheng, Yanbing Ma, Jianrong Zhou, Weiding Zhai, Binbin Su, Shoujun Li, Li Chen

## Abstract

**Background:** Current circulating biomarkers for idiopathic pulmonary arterial hypertension (IPAH) lack specificity for preclinical detection and fail to capture the biological heterogeneity driving disease progression. Furthermore, molecular mechanisms underlying the “sex paradox” of IPAH, where females exhibit higher susceptibility but lower mortality, remain poorly understood, hindering the development of precision therapeutics.

**Methods:** We performed an integrated proteogenomic analysis characterizing 2,920 plasma proteins from 45,811 participants in the UK Biobank. We integrated discovery-driven Cox regression with case-control verification, followed by cis-Mendelian randomization and colocalization to distinguish causal mediators from bystanders. We utilized unsupervised clustering for biological risk stratification and machine learning to deconvolute sex-dimorphic proteomic signatures, finally applying systemic drug reproposing to prioritize therapeutic candidates.

**Results:** We identified 92 causal proteins driving IPAH incidence (e.g., NOTCH3, FLT3IG) and 15 driving mortality (e.g., REG4, CA6), with 9 proteins (e.g., EDN1, LRRN1) serving as dual determinants. Unsupervised clustering identified a high mortality-risk phenotype characterized by upregulated proteins associated with extracellular matrix-receptor interaction, transforming growth factor-β signaling, cardiac hypertrophy, and elastic fibre formation, together with reduced plasma levels of APOL1. Notably, sex-stratified analysis revealed divergent pathogenic architectures that progression of IPAH in males was predominantly linked to right ventricular dysfunction mediators (e.g., NT-proBNP, GDF15), whereas in females, it was more strongly driven by vascular dysfunction mediators (e.g., EDN1, BCL2L15). Finally, we prioritized 27 druggable targets, with genetic evidence highlighting AGRN, CLU, and DDR1 as high-confidence candidates for therapeutic intervention.

**Conclusions:** This study delineates causal proteomic landscape of IPAH, bridging epidemiological associations with genetically supported targets. By uncovering the molecular basis of sex-dependent outcomes and prioritizing novel druggable proteins, our findings provide a robust framework for preclinical detection, innovative risk stratification, and the development of precision therapeutics.

Central Abstract.
A causal proteogenomic atlas for idiopathic pulmonary arterial hypertension

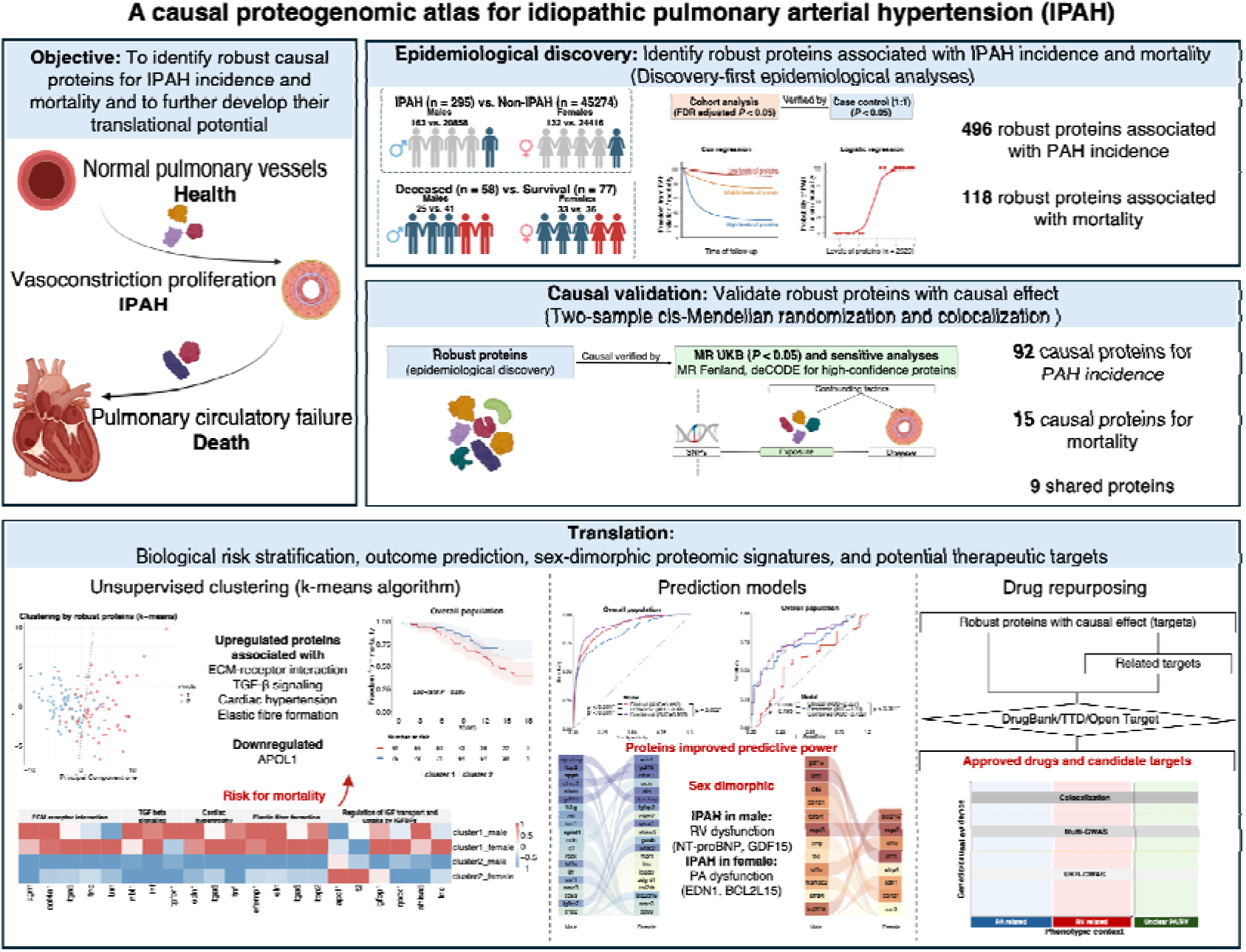

## Introduction

Idiopathic pulmonary arterial hypertension (IPAH) is a rare but devastating cardiovascular disease, characterized by a progressive increase in pulmonary vascular resistance of largely undetermined origin and the subsequent development of right ventricular failure ^1^. Despite therapeutic advances, the prognosis remains poor, with a case-fatality rate exceeding 10% annually ^2^. Current guidelines underscores circulating biomarkers as indispensable tools for prognostic assessment ^3,4^. B-type natriuretic peptide (BNP) and N-terminal prohormone of BNP (NT-proBNP) represent the biomarkers routinely employed for risk stratification in clinical practice ^3,4^. However, their utility is constrained by suboptimal specificity and considerable susceptibility to confounding, such as age, obesity, inflammation, renal function, and cardiovascular comorbidities ^5–9^. Moreover, as myocardial strain-responsive alarms rather than primary drivers of pulmonary vascular remodeling^10^, their indirect reflection and intrinsic temporal delay to vascular pathology hinder their utility of preclinical detection ^10^.

Circulating proteins, as direct effectors of intercellular signaling, offer a dynamic window into the active processes of pulmonary vascular constriction, dysfunction, and proliferation ^11^. However, translating proteomic signatures into clinical utility has been hindered by methodological constraints. Most prior studies were limited to conventional epidemiological associations within modest and disease-specific cohort, posing two critical challenges ^12–15^. First, it is difficult to distinguish causal drivers of disease onset from downstream bystanders of established cardiac dysfunction. Second, it fails to capture the preclinical status, leaving the early determinants of IPAH incidence largely unknown. Furthermore, primary IPAH exhibits a profound epidemiological sex dimorphism, where females present higher disease susceptibility but better survival than males^16,17^, yet the molecular underpinnings of this paradox remain poorly characterized.

Addressing these gaps requires a transition from single observational studies to causally anchored biological inferences. A systematic integration of prospective epidemiology with genetic instrumentation provides a robust framework to filter out confounders and identify causal proteins with translational potential ^18^. Here, we present a systematic proteomic investigation leveraging a large-scale, prospective population-based cohort to interrogate the biological architecture of IPAH. We designed a coherent analytical framework integrating discovery-driven epidemiology, multi-design replication, and cis-MR. Our objectives were to determine robust causal proteins for IPAH incidence and mortality, delineate distinct high-risk biological endotypes, understanding sex-dimorphic proteomic signatures, and accelerate the identification of therapeutic targets.

## Results

### Study population

A total of 45,569 participants without a diagnosis of IPAH at baseline comprised the incidence cohort and 135 participants diagnosed with IPAH at baseline formed the mortality cohort. During a median follow-up of 15 years, 295 of 45569 participants (0.6%) in the incidence cohort were newly diagnosed with IPAH and 58 of 135 patients with IPAH (43%) in the mortality cohort died. To comprehensively identify proteins associated with incidence and mortality, participants in each cohort were pre-stratified by sex to mitigate the potential impact of sex-dependent heterogeneity.

### Robust proteins associated with IPAH incidence and mortality

In a pre-specified, sex-stratified prospective cohort, we used a discovery-first strategy with a minimal sufficient covariate set and FDR control, then strengthened signals by case-control verification. This strategy was designed to yield robust and cross-design consensus protein sets that anchor subsequent genetic validation and translational analyses. (Fig.2A)

**Figure 1.**
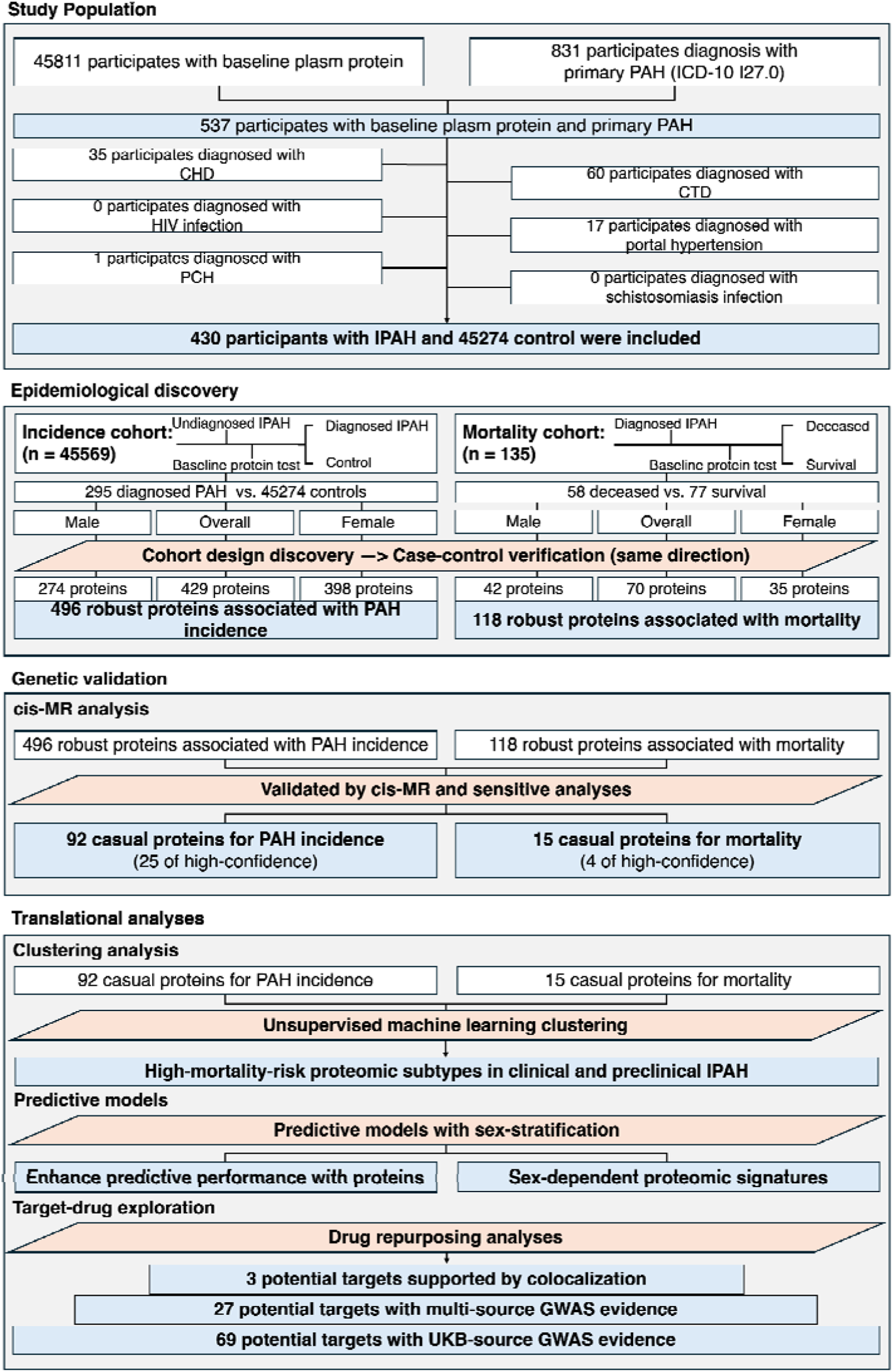
Study design. Abbreviations: PAH, pulmonary arterial hypertension; CHD, congenital heart disease; CTD, connective tissue disease; HIV, human immunodeficiency virus; PCH, pulmonary capillary hemangiomatosis; IPAH, idiopathic pulmonary arterial hypertension; MR, mendelian randomization.

**Figure 2.**
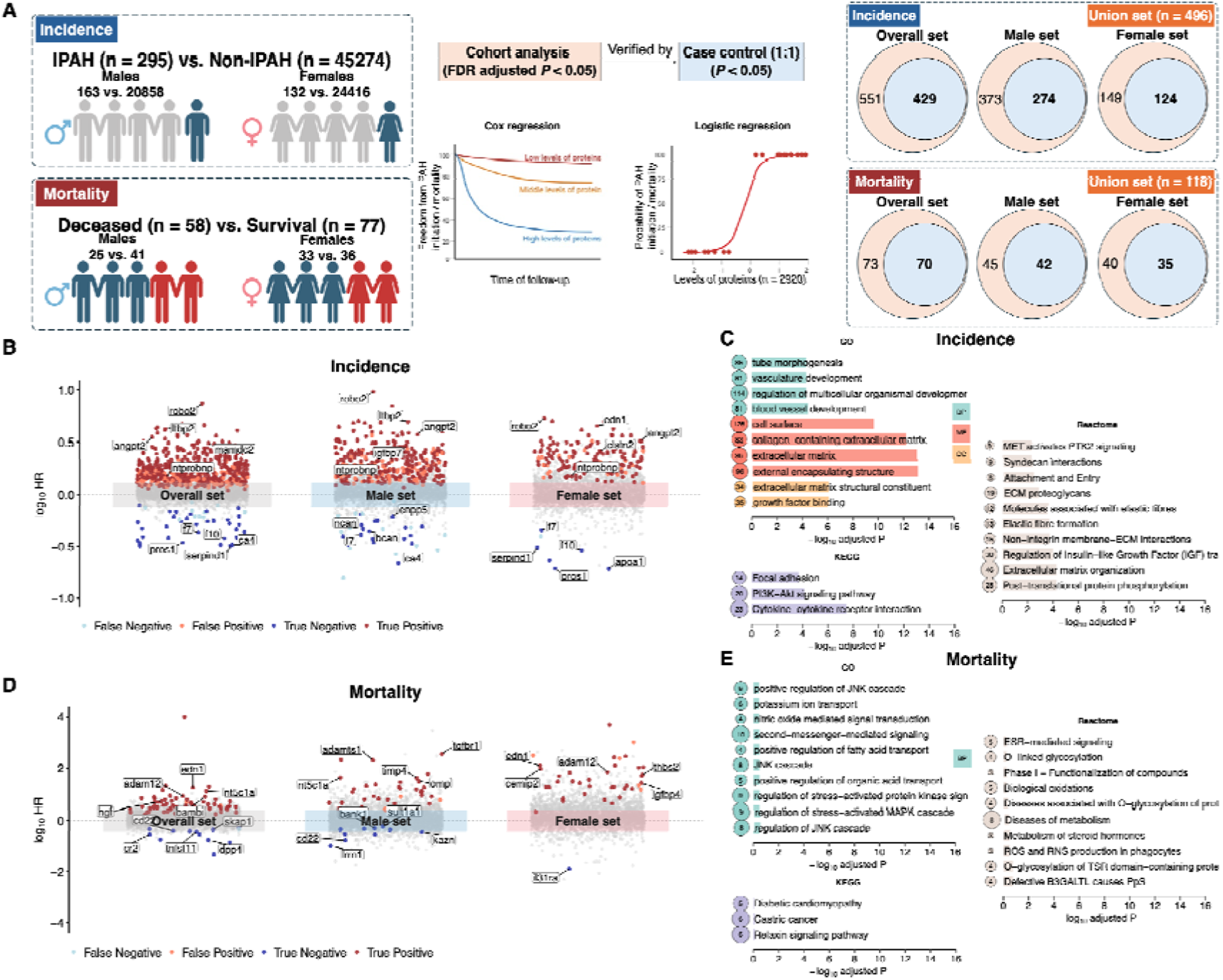
Discovery of robust proteins associated with IPAH incidence and mortality using epidemiological analyses. **Notes: (A)** Design of the discovery process for robust proteins associated with IPAH incidence and mortality **(B)** Robust proteins associated with incidence. This plot displays the effect size using log_10_(HR) in cox regression models. Proteins with statistical significance (FDR-adjusted *p*-value < 0.05 in the cohort design and nominal *p*-value < 0.05 in the case–control design) and concordant effects in both designs were defined as true positives or true negatives, while proteins showing statistical significance and opposite directions of effect relative to the cohort design were classified as false positives or false negatives. **(C)** Enrichment analyses for robust proteins associated with incidence (n = 496, background = 2920), with top 10 GO terms, top 3 KEGG pathways, and to 10 Reactome pathways ranked by significance are shown. **(D)** Robust proteins associations with mortality. This plot displays the effect size using log_10_(HR) in cox regression models. Proteins with statistical significance (FDR-adjusted *p*-value < 0.05 in the cohort design and nominal *p*-value < 0.05 in the case–control design) and concordant effects in both designs were defined as true positives or true negatives, while proteins showing statistical significance and opposite directions of effect relative to the cohort design were classified as false positives or false negatives. **(E)** Enrichment analyses for robust proteins associated with mortality (n = 118, background = 2920), with top 10 GO terms, top 3 KEGG pathways, and top 10 Reactome pathways ranked by significance are shown. Abbreviations: PAH, pulmonary arterial hypertension; FDR, false discovery rate; HR, hazard ratio; GO, gene ontology; BP, biological process; MF, molecular function; CC, cellular component; KEGG, Kyoto encyclopedia of genes and genomes.

In the observation of incidence cohort, discovery identified 551 proteins associated with IPAH incidence, with 373 proteins in males and 149 proteins in females. Association remained after case-control verification, resulting an incidence-consensus set of 496 proteins (429 of overall, 274 of males, and 127 of females). Among robust proteins validated by case-control design, 454 showed a positive association, whereas 42 proteins exhibited negatively. These proteins were enriched for extracellular matrix (ECM) organization, vascular and tube morphogenesis, and growth factor binding; the KEGG and Reactome highlighted PI3K-Akt signaling, cytokine-receptor interaction, and elastic fiber formation. (Fig.2B-C)

In the mortality cohort, discovery yielded 73 mortality-associated proteins, with 45 in males and 40 in females. Signals persisted under case-control verification, defining a mortality consensus set of 118 proteins (70 of overall, 42 of males, and 35 of females) that enriched in metabolic processes, oxidative stress response, and JNK/MAPK signaling pathways. (Fig.2D-E)

### Causal prioritization of proteins

Next, we estimated the causal effect of these robust proteins for IPAH incidence and mortality using two-sample cis-MR and sensitivity analyses. Also, we performed colocalization to determine whether causal proteins and IPAH phenotype have shared causal variant. (Fig.3A)

**Figure 3.**
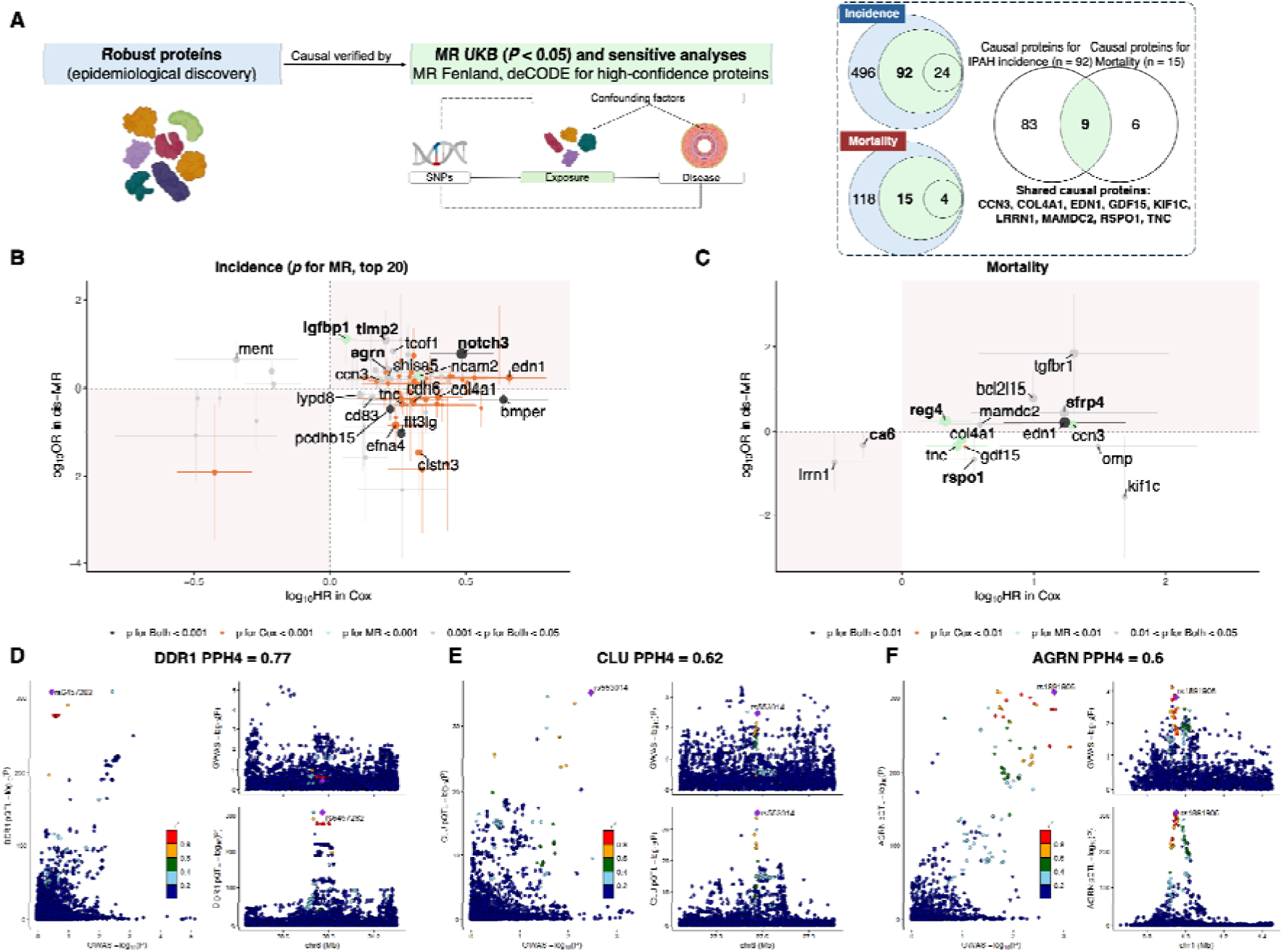
Causal effect validation of robust proteins for IPAH incidence and mortality using cis-MR, sensitive analyses, and colocalization. **Notes: (A)** Design of validation process for causal effect of robust proteins for IPAH incidence and mortality. In the incidence cohort, 92 robust proteins were supported by UKB MR and 24 of total were defined as high-confidence supported by multi-source MR; In the mortality cohort, 15 robust proteins were supported by UKB MR and 4 of total were defined as high-confidence supported by multi-source MR; 9 proteins (i.e., CCN3, COL4A1, EDN1, GDF15, KIF1C, LRRN1, MAMDC2, RSPO1, TNC) were shared by IPAH incidence and mortality. **(B)** Comparison of effective sizes associated with incidence in epidemiological analyses (log_10_HR in cox regression model, x-axis) and cis-MR analyses (log_10_OR in cis-MR, y-axis). Proteins in the upper-right and lower-left quadrants show concordant directions, with dot colors distinguishing statistical significance in the discovery and validation outcomes. Top 20 significant proteins in cis-MR results are presented and highlighted in bold are high-confidence. **(C)** Comparison of effective sizes associated with mortality in epidemiological analyses (log_10_HR in cox regression model, x-axis) and cis-MR analyses (log_10_OR in cis-MR, y-axis) for mortality. Proteins in the upper-right and lower-left quadrants show concordant directions, with dot colors distinguishing statistical significance in the discovery and validation outcomes. Proteins highlighted in bold are high-confidence. **(D)** Regional colocalization for DDR1 with IPAH. **(E)** Regional colocalization for CLU with IPAH. **(F)** Regional colocalization for AGRN with IPAH Abbreviations: FDR, false discovery rate; MR, mendelian randomization; UKB; UK biobank; IPAH, idiopathic pulmonary arterial hypertension; HR, hazard ratio; OR, odds ratio.

Among robust proteins associated with incidence, 92 causal proteins (24 high-confidence) were confirmed, with 44 proteins showing consistently positive effects and 5 showing consistently negative effects in both epidemiological and cis-MR analyses. Notably, NOTCH3 (*p*-value for both < 0.001), IGFBP1 (*p*-value for MR < 0.001), and NCAM2 (*p*-value for MR < 0.001) presented the top 3 significant positive proteins. Conversely, other 43 proteins exhibited opposing effect direction, with FLT3IG (*p*-value for both < 0.001), PCDHB15 (*p*-value for both < 0.001), and BMPER (*p*-value for both < 0.001) being the most significant proteins in cis-MR analyses. (Fig.3B) For robust proteins in mortality cohort, 15 (4 of high-confidence) proteins with causal effect were confirmed. Therein, 7 exhibited consistent positive effects, 2 demonstrated consistent negative effects, and 6 showed opposing effects. Endothelin-1 (EDN1) was the only protein with a *p*-value < 0.01 for both epidemiological and cis-MR analyses. (Fig.3C) Robust proteins demonstrated causal effects were then employed in downstream analyses to explore their translational potential. Colocalization-based causal prioritization identified Epithelial discoidin domain-containing receptor-1 (DDR1) (PPH4 = 0.77), Clusterin (CLU) (PPH4 = 0.62), and Agrin (AGRN) (PPH4 = 0.6) as the proteins likely driven by a shared genetic variation with IPAH phenotype, prioritizing it for subsequent validation. (Fig.3D-F)

### Proteomic subtypes and biological risk stratification

Robust proteins with causal effect, identified through an integrated framework of epidemiological discovery and genetic validation, served as a promising instrument for biological risk stratification. We then performed clustering analyses to identify proteomic-defined high-risk subtypes for mortality.

Patients with IPAH at baseline were grouped into two proteomic subtypes. In both the overall cohort and among male patients, those with a proteomic profile characterized by elevated circulating proteins involved in extracellular matrix–receptor interaction, TGF-β signaling, cardiac hypertrophy, and elastic fibre formation, together with reduced plasma levels of APOL1, had a higher risk of mortality than those with the contrasting proteomic signature. However, this association was not significant in female patients. (Fig.4A-C) In a complementary analysis of the preclinical IPAH incidence cohort, individuals assigned to the corresponding proteomic subtype exhibited similar high-risk pathway patterns for mortality irrespective of sex. In addition, overall mortality was higher among preclinical IPAH cases, individuals who were free of IPAH at baseline but subsequently developed the disease, than among patients with established IPAH at baseline. (Fig.4D-F) Proteomic signatures for clinical patients with IPAH and preclinical patients at individual level were shown stratified by clusters and sexes in the Supplementary Fig.1 and Fig.2, respectively.

**Figure 4.**
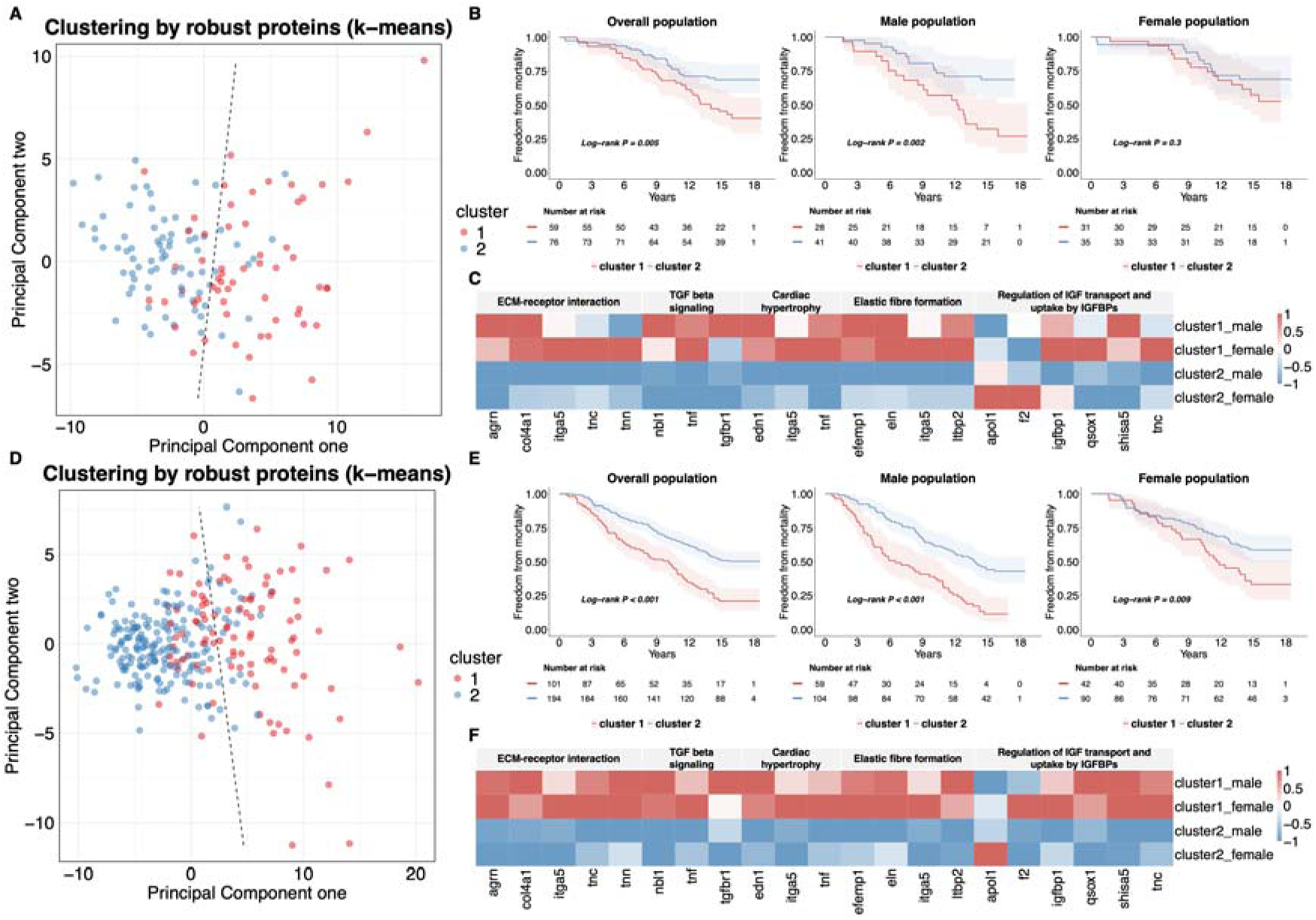
Biological risk stratification for mortality using clustering analyses. **Notes: (A)** PCA-based k-means clustering for clinical patients with IPAH. **(B)** Freedom from mortality for clinical patients with IPAH grouped by proteomic clusters and sex-stratified subgroup. **(C)** Sex-and cluster-stratified heatmap of mean protein z-scores. Standardized z-scores were truncated to the range −1 to 1, with higher expression (z closer to 1) shown in red and lower expression (z closer to −1) shown in blue. **(D)** PCA-based k-means clustering for preclinical patients without IPAH. **(E)** Freedom from mortality for preclinical patients without IPAH grouped by proteomic clusters and sex-stratified subgroup. **(F)** Sex-and cluster-stratified heatmap of mean protein z-scores. Standardized z-scores were truncated to the range −1 to 1, with higher expression (z closer to 1) shown in red and lower expression (z closer to −1) shown in blue. Abbreviations: ECM, extracellular matrix; TGF, transforming growth factor; IGF, insulin-like growth factor; IGFBPs, insulin-like growth factor binding proteins; PCA, principal component analysis; IPAH, idiopathic pulmonary arterial hypertension;

### Risk prediction and sex-difference of casual proteins for IPAH incidence and mortality

Robust proteins with causal effect were then incorporated into the predictive model to examine their potential contribution to predictive performance for IPAH incidence and mortality. In terms of predicting incidence, although proteomic model exhibited a lower predictive performance, integrating proteins with clinical factors enhanced the overall predictive power (combined AUC: 0.909 vs. clinical 0.887, *p*-value = 0.002). (Fig.5A) Also, proteomic predictors outperformed clinical variables in mortality prediction, improving AUC from 0.557 of clinical model to 0.762 of combined model in overall population (*p*-value < 0.001) and from 0.675 to 0.822 in males (*p-*value = 0.041). (Fig.5D)

**Figure 5.**
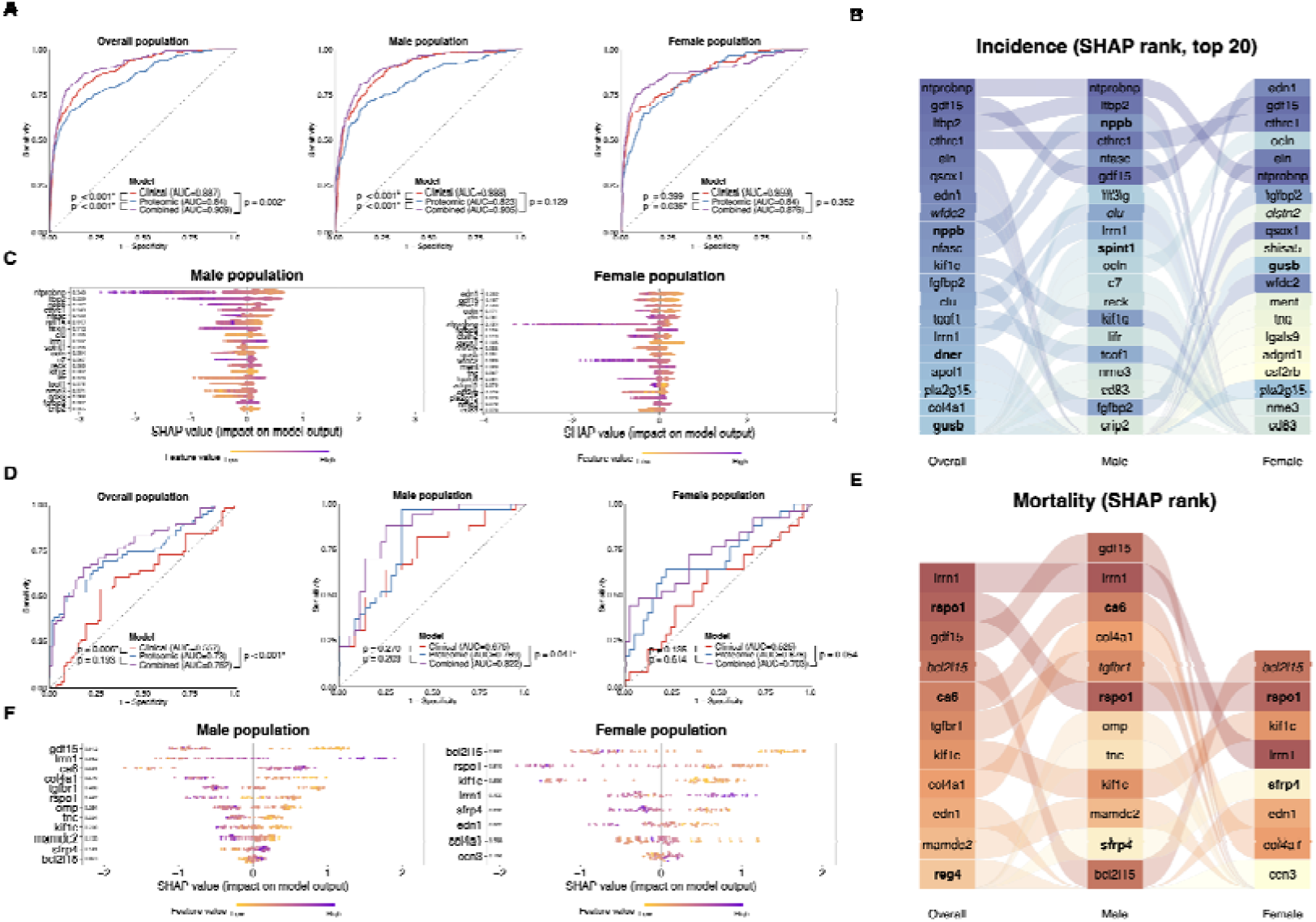
Risk prediction and sex-dimorphic proteins for IPAH incidence and mortality. **Notes: (A)** Performance of prediction models for incidence across overall, male, and female population. **(B)** SHAP rank of predictive proteins for IPAH incidence. Sankey plot presents the top 20 predictive proteins for incidence across overall, male, and female population and high-confidence proteins were bold. **(C)** SHAP value of predictive proteins for IPAH incidence between male and female population. **(D)** Performance of prediction models for mortality across overall, male, and female population. **(E)** SHAP rank of predictive proteins for mortality. Sankey plot presents all predictive proteins for mortality across overall, male, and female population and high-confidence proteins were bold. **(F)** SHAP value of predictive proteins for mortality between male and female population. Abbreviations: AUC, area under the curve; IPAH, idiopathic pulmonary arterial hypertension; SHAP, SHapley Additive exPlanations.

Considering the sex-related differences in IPAH incidence and mortality potentially driven by robust proteins with causal effect, we assessed their contribution in prediction models for sex-stratified subgroups by ranking them with SHAP value. Results indicated NTPROBNP, GDF15, and CTHRC1 stably contributed to incidence for both sexes. However, remarkable sex-dependent differences remained, with LTBP2, NPPB, and NFASC pronounced in males but EDN1, ELN, and FGBP2 striking in females. (Fig.5B-C) Sex-specific proteomic patterns also observed in the mortality cohort, with GDF15, CA6, TGFBR1, OMP, TNC, and MAMDC2 predicting mortality exclusively in males, whereas EDN1 and CCN3 predicted mortality only in females. (Fig.5E-F)

### Therapeutic targets and drug repurposing opportunities

A total of 98 potential therapeutic targets for IPAH were identified, of which 27 were classified as high-confidence and 3 were supported by colocalization. (Figure 6) To access potential therapeutic value for these three tired robust proteins, we systematically summarized proteomic targets and their related targets using DrugBank, the Therapeutic Target Database, and the Open Targets database. Among all druggable targets, 28 had supporting evidence for involvement in vascular impairment, and 8 had evidence linking them to cardiac dysfunction. Among targets with approved drugs, EDN1 is an acknowledged pathogenic protein of pulmonary artery hypertension (PAH), with its receptor ETA recognized as the one of the most significant therapeutic targets. Bosentan and its analogs act by blocking ETA to mitigate vasoconstriction and vascular remodeling during disease progression. Moreover, GDF11, a member of the TGF-β superfamily, signals through TGFBR1, which was identified as a robust risk protein for IPAH incidence. Sotatercept, a ligand trap targeting multiple TGF-β superfamily members including GDF11, modulates this pathway and thereby inhibits vascular muscle cell proliferation. Additional proteins and related targets uncovered through the same process may be higher druggable and could serve as potential candidates for future therapeutic development. (Supplementary Table 1)

**Figure 6.**
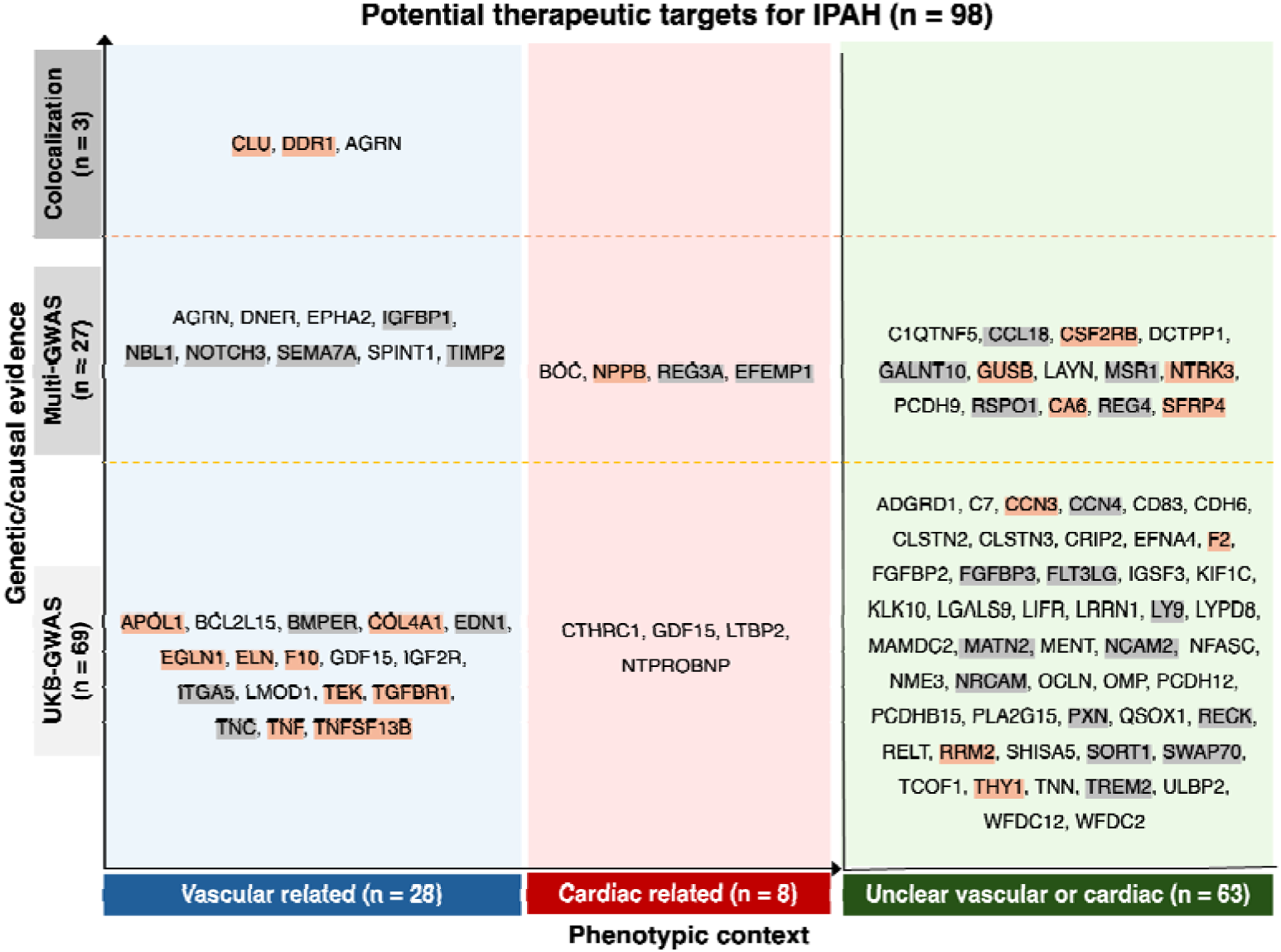
Integrated prioritization of potential therapeutic targets for IPAH. **Notes:** 98 proteins with putative causal effects on IPAH were mapped in a two-dimensional framework according to the strength of genetic/causal evidence (y-axis) and phenotypic context (x-axis). The y-axis distinguishes targets supported only by UKB-GWAS evidence (n = 69), those additionally replicated in external multi-GWAS evidence (n = 27), and those supported by colocalization (n = 3). The x-axis separates targets primarily related to vascular impairment (left in blue, n = 28), cardiac dysfunction (middle in red, n = 8), and those with unclear specificity (right in green, n = 63). Orange labels denote targets with drugs directly approved against them, grey labels indicate targets with approved drugs on related pathways, and the remaining targets currently have no approved therapies.

## Discussion

IPAH is a rare but fatal condition, and risk stratification and therapeutic targets exploration are essential to improving prognosis. Here, leveraging a proteogenomic atlas derived from 2,920 plasm proteins of 430 patients diagnosed IPAH and over 450,000 controls, we defined two panels of robust causal proteins that may facilitate identifying mortality-prone proteomic phenotype, informing sex-dimorphic pathogenic differences, and accelerating therapeutic medication development for IPAH. The coherent research framework constitutes a major strength of this study that interpreting proteome-wide association, methodological replication, genetic validation of MR to discovery high-confidence proteins and exploring the translational potential using unsupervised clustering, prediction model, and drug repurposing.

Progression to secondary right ventricular (RV) dysfunction remains the key pathophysiological determinant driving the clinical manifestation and prognosis in patients with IPAH ^1^. Accordingly, the 2022 ESC/ERS guidelines recommend BNP and NT-proBNP as the sole biomarkers for clinical risk stratification ^3^. Our results corroborate this established clinical paradigm, demonstrating that preclinical elevations in BNP and NT-proBNP are positively associated with subsequent IPAH onset, likely reflecting subclinical RV overload. Significantly, other cardiac related biomarkers, such as BOC ^19^, REG3A ^20^, EFEMP1 ^20^, CTHRC1 ^21^, and LTBP2 ^22^ have emerged as risk proteins specifically linked to RV dysfunction due to pulmonary hypertension, reflecting their effective ability to capture hemodynamic consequence. Our findings further substantiate their value by highlighting its predictive capacity for disease onset. Additionally, biomarkers reflecting the underlying pulmonary vascular pathology remain essential for elucidating disease mechanisms and achieving early detection. EDN1 is a canonical protein upregulated in response to endothelial injury and dysfunction ^23^, was reaffirmed in this study as a causal contributor to both disease development and mortality. Consistent with mechanistic data showing that EPHA2 deficiency aggravates endothelial dysfunction and PAH severity ^24^, we found elevated higher preclinical circulating EPHA2 was associated with an increased risk of IPAH onset. Also, CLU ^25^, IGFBP1 ^26^, and NOTCH3 ^27,28^ signaling is known to promote pulmonary artery smooth muscle cells (PASMCs) proliferation in PAH and our data adds clinical evidence that these proteins are elevated prior to diagnosis, thereby supporting its potential as an early biomarker of vascular remodeling. Collectively, proteins identified within our analytical framework serve as robust circulating biomarkers for PAH onset or progression and their consistency with previous evidence reinforces the robustness and credibility of our findings.

In addition to these established biomarkers, we further confirmed a panel of exploratory circulating proteins with the potential to be involve in IPAH progression and prognosis. AGRN, originally recognized for its critical role in neuromuscular junction formation, has been found to be implicated in tumor angiogenesis ^29^ and heart regeneration ^30^ and is overexpressed within the ECM of PAH lung tissue ^31^. Together, the AGRN may contribute to the establishment of a tumor-like ECM niche in pulmonary arterial wall, thereby prompting vascular remodeling, although this warrants1 further mechanistic validation. EFEMP1, an ECM glycoprotein that regulates cell adhesion and migration through epidermal growth factor receptor signaling, and REG3A, a secreted C-type lectin implicated in innate immunity and tissue repair, have both been associated with RV dysfunction in PAH, yet their mechanistic contributions to PAH progression remain unclear ^20^. In brief, the identification of these putative proteomic biomarkers underscores the complexity of PAH and provide compelling evidence for functional investigations and translational biomarker strategies.

Integration of these two iteratively derived protein panels showed strong performance in discriminating high-from low-mortality risk patients across both the clinical and preclinical IPAH cohorts. Indeed, as previous studies, biological stratification using circulating proteins helps distinguish patients with PAH or pulmonary hypertension at higher risk of poor prognosis ^13,15^. J Sweatt et al. identified immune phenotype profiled with elevated circulating tumor necrosis factor-related apoptosis-inducing ligand, C-C motif chemokine ligand 5 (CCL5), CCL7, CCL4, and macrophage migration inhibitory factor, was associated with higher mortality risk among patients with PAH ^13^. Boucly et al. applied unsupervised clustering to 156 plasm proteins and identified a subgroup of pulmonary hypertension patients marked by an upregulated Transforming Growth Factor-β (TGF-β) pathway who subsequently experienced poorer clinical outcomes ^15^. Our data provide converging evidence that patients with a proteomic profile enriched for elevated proteins involved in ECM-receptor interaction, TGF-β signaling, cardiac hypertrophy, and elastic fibre formation constitute a biological phenotype at substantially increased risk of mortality, with this association being particularly pronounced in males. Emerging data indicate that the male RV is less capable of maintaining adaptive remodeling under chronic pressure overload and is more prone to maladaptive deformation and dysfunction ^32^. In this context, a proteomic pattern in which ECM and PASMCs remodeling act as the initiating insult and RV hypertrophy and fibrosis represent downstream consequences is likely to be amplified in men, thereby further escalating their risk of death. Additionally, APOL1, an HDL-associated, inflammation-inducible apolipoprotein best known as a major “kidney-risk” gene ^33^, has not previously been implicated in IPAH, yet its roles in endothelial injury provide a biologically plausible link to vascular remodeling ^34^. Here, however, lower circulating APOL1 levels partially characterized the mortality-prone proteomic subtype, suggesting a complex, context-dependent relationship between APOL1 signaling and disease progression that warrants rigorous mechanistic dissection.

Consistent with previous studies, risk-associated proteins offer independent prognostic value and can also incrementally enhance clinical assessments for mortality prediction. J Rhodes et al. demonstrated a panel of nine-proteins could identify PAH patients at high-risk of mortality without clinical evaluation ^12^, while a minimal six-proteins subset further improved transplantation-free survival prediction when combined with NT-proBNP ^35^. In our study, although we employed a filtered set of mortality-associated proteins in PAH patients, predictive performance for mortality was slightly inferior to prior reports ^12,35^. This is plausibly due to the inability to incorporate established clinical risk scores, such as REVEAL, within a natural population framework. Crucially, however, assessment tools like REVEAL depend heavily on invasive hemodynamics obtained via right heart catheterization, as well as clinician-dependent subjective assessments such as WHO functional class ^16^. By achieving robust discrimination using plasma proteins alone, our models demonstrate a distinct translational advantage that consists in their use as a non-invasive screening tool suitable for primary care or resource and resource-limited settings.

In our study, the primary purpose of our predictive models was not designed to improve predictive performance, but to understand sex-dimorphism in the roles of individual proteins in driving disease development and mortality. Epidemiologically, PAH demonstrated a pronounced sex paradox, with females disproportionately affected by disease onset ^16^, while males exhibiting a higher risk of death ^17^. From large-scale circulating proteome, we inform novel insights regarding on this sex divergence of disease onset that RV dysfunction associated proteins, such as NT-proBNP of incidence and GDF15 of mortality ranked the top contributors in males, whereas vasoconstriction related proteins, such as EDN1 of incidence, contributed prominently to females. Estrogens exert a vasculoprotective influence in part by suppressing EDN1 gene expression and lowering circulating EDN1 ^36^. Perimenopausal women experience pronounced fluctuations and overall decline in estrogen levels, and the resulting instability and attenuation of estrogen-mediated vascular protection may indirectly enhance the biological effects of EDN1, thereby amplifying its pathogenetic role in PAH. Moreover, animal models have shown that testosterone promotes maladaptive RV hypertrophy and fibrosis under pressure overload ^37^. Consistently, a population-based observational study demonstrated that men have greater RV mass, reduced RV ejection fraction, and poorer RV-pulmonary artery coupling than women ^32^. Insightfully, hormones are not the sole drivers for sex dimorphism. An in-vitro study using shear-stress-treated human pulmonary microvascular endothelia cells demonstrated inherent sex divergence in both RNA and protein expression, with functional manifestation of higher proliferative rate in male cells in normoxic environment relative to female cells. Our findings further revealed that proteins implicated in PASMC proliferation (e.g., TGFBR1) and ECM remodelling (e.g., TNC) contributed more substantially to mortality risk in men, whereas the vasoconstriction-related mediator EDN1 remained exert a greater impact in women. Collectively, the pronounced sex dimorphism in causal proteins supports the presence of sex-specific biological pathways in the development and prognosis of IPAH and may inform more tailored prevention and treatment strategies.

Targeted drug treatment represented the cornerstone of long-term IPAH management, focusing on three key pathways, EDN1 signalling (endothelin-receptor antagonists), the nitric oxide–cGMP pathway (PDE5 inhibitors and soluble guanylate cyclase stimulators), the prostacyclin pathway (prostacyclin analogues and IP-receptor agonists) ^3,4^. More recently, agents targeting the BMPR2-TGF-β signalling axis (growth differentiation factor modulators) have emerged as a promising fourth therapeutic pathway in IPAH ^38^. Here, coherent identification of robust protein targets recapitulated two of these established therapeutic pathways, the EDN1 and BMPR2–TGF-β signalling axis, providing confidence to further explore the therapeutic potential of other identified protein targets. Among these potential targets, the ECM secreted protein AGRN emerged as the pinnacle of our tripartite stratification hierarchy; however, no approved drugs currently act on this molecule, highlighting it as a promising yet undeveloped therapeutic opportunity. Furthermore, our data highlighted two additional robust targets of interest using colocalization analyses. CLU is a secreted glycoprotein that has been identified as overexpressed in lung tissue and implicated in promoting PASMCs proliferation, migration, and resistance to apoptosis in preclinical models ^25^. Moreover, as a metalloprotein, CLU’s structural and functional activities are regulated by zinc and copper, positioning these metal ions as potential therapeutic regulators in the context of PAH. DDR1 functions as an extensively expressed collagen-binding receptor tyrosine kinase (RTK) involved in ECM remodeling. Our cis-MR analysis identified a robust positive causal effect of DDR1 on IPAH, implicating elevated DDR1 signaling as a pathogenic driver. This human genetic evidence offers a distinct perspective from previous murine studies, where global DDR1 deletion was reported to exacerbate vascular remodeling ^39^, suggesting significant species-specific or context-dependent mechanisms. Translating our finding into therapy presents a unique pharmacological challenge. Though DDR1 inhibition is the logical strategy, potent DDR1 inhibitor Dasatinib is clinically contraindicated due to its association with drug-induced PAH ^40^. This toxicity is likely attributable to off-target inhibition RTK rather than DDR1 blockade itself. Therefore, our results do not support the use of existing broad-spectrum RTK inhibitors but instead highlight a critical need for highly selective DDR1 inhibitors. Such tools are essential to resolve the biological directionality of DDR1 in humans and decouple therapeutic benefit from the vascular toxicity of promiscuous agents.

Several inherent limitations of our study should be addressed. First, although a sequential design was implemented to strengthen internal validity, the absence of an independent external cohort with similar proteomic and phenotypic depth hinders the assessment of generalizability across diverse populations and ethnic groups. Second, the use of ICD-10 codes I27.0 for IPAH case identification in natural population-based cohorts may have introduced misclassification and subsequent selection bias, for example by inadvertently including cases of secondary pulmonary hypertension or failing to capture true IPAH. Third, inherent constraints of the natural population-based cohort limited the availability of key clinical covariates, such as direct assessments of hemodynamics, RV function, and exercise capacity, preventing full adjustment for established clinical predictors and necessitating validation of our results in disease-specific cohorts. Last, from a translational standpoint, targeted therapies for key candidate proteins remain underdeveloped, and the low utilization of existing pathway-specific agents in the cohort limited any robust pharmacoepidemiologic or real-world evidence analyses.

In summary, this study delineates both preclinical and clinical high-resolution causal proteomic atlases of IPAH, bridging the critical gap between genetic susceptibility and clinical manifestation. Our transition from relying on adaptive hemodynamic biomarkers to identifying preclinical drivers provides a robust framework for early detection in the general population. Also, our novel molecular stratification has identified a high-risk mortality subgroup that predominantly male-dependent. Furthermore, by deconvoluting the molecular architecture of the “sex paradox”, we reveal that male and female disease trajectories are governed by divergent biological imperatives, cardiac failure versus vascular regulation, necessitating a shift towards sex-dimorphic precision medicine. Collectively, these findings move the field beyond descriptive associations, offering a genetically anchored roadmap for developing the precision therapeutics.

## Method

### Study population and grouping strategy

The UK Biobank (UKB) is a large-scale, population-based prospective cohort study that recruited over 500,000 participants aged 40 to 69 years from 22 assessment centers across the United Kingdom between 2006 and 2010. Here, we leveraged demographic, diagnostic, laboratory, plasm proteomic, and genetic data from 45,811 randomized UKB participants, comprising 537 patients with pulmonary arterial hypertension. To further restrict to the IPAH, we excluded other secondary PAH with the following comorbidities: congenital heart disease (n = 35), connective tissue disease (n = 60), human immunodeficiency virus infection (n = 0), portal hypertension (n = 17), pulmonary capillary hemangiomatosis (n = 1), or schistosomiasis (n = 0). Ultimately, a total of 430 participants with IPAH and 45,274 without the disease were retained. Additionally, to assess proteomic profiles associated with disease stage, participants were further divided into PAH incidence cohort and mortality cohort based on the temporal relationship between enrollment and the date of diagnosis. In particular, the PAH incidence cohort comprised 295 participants diagnosed IPAH after baseline assessment and compared with those without IPAH diagnosis; while the mortality cohort included 135 participants diagnosed IPAH before baseline assessment and follow-up to observe all-cause mortality. (Fig.1) The UK Biobank received ethical approval from the Northwest Multi-center Research Ethics Committee (11/NW/0382), and all participants provided written informed consent. This study was conducted under UK Biobank application number 105435.

### Proteomic measurement and data process

EDTA-plasma samples were collected from 53,004 UKB participants between 2006 and 2010, representing a randomized subset of the UK Biobank Pharma Proteomics Project (UKB-PPP) Consortium. Samples were stored at -80[°C and transported on dry ice to the Olink Explore 1536 and Expansion platforms for proteomic profiling. A total of 2,923 unique proteins were quantified across 2,941 assays, arranged in four 384-plex panels. Protein abundances were expressed as log2-transformed Normalized Protein eXpression (NPX) values to enable standardized comparison. Participants with over 50% missing protein data, three proteins missing in more than half of the cohort, and participants enrolled to drug trails were excluded. After quality control, 2920 proteins across 45,811 participants were retained for downstream analyses, yielding a robust dataset for high-resolution proteomic investigation. NPX values with missing entries were imputed via a chained random forests algorithm incorporating age and sex as predictors and subsequently standardized to a standard normal distribution with a mean of 0 and a standard deviation of 1.

## Definition

### Pulmonary arterial hypertension

Diagnosed PAH was identified from linked hospital inpatient and primary care data from the UKB, corresponding to the International Classification of Disease, 10^th^ Revision (ICD-10) code I27.0 for primary PAH. To confine the cohort to IPAH, cases diagnosed with following associated conditions was excluded ^41^.

Congenital heart disease: Q20.0 to Q20.5, Q25.0, Q21.x, Q26.2, and Q26.3

Connective tissue disease: L90.0, L94.0, L93.x, M05.x, M06.x, M08.x, M12.0xx, M32.x, M33.x, M34.x, M35.0x, M35.1, M35.8, and M35.9

Human immunodeficiency virus infection: B20.x, B21.x, B22.x, B23.x, B24.x, B97.35, Z21.x

Portal hypertension: K76.6

Pulmonary capillary hemangiomatosis: I78.0

Schistosomiasis infection: B65.x

### Covariates

Covariates were obtained from the UKB baseline assessments. Demographic and clinical variables included age (continuous, years), sex (binary, male or female), ethnicity (binary, white or others), body mass index (BMI; continuous, kg/m²), systolic blood pressure (continuous, mmHg), and diastolic blood pressure (continuous, mmHg). Lifestyle factors comprised smoke status (binary, yes [current/former] or no) and alcohol status (binary, yes [current/previous] or no). Comorbid cardiovascular diseases were identified using ICD-10 codes ^42^, including hypertensive disease (binary, yes [I10-I15] or no), mellitus (binary, yes [E10-E14] or no), coronary artery disease (binary, yes [I25] or no), and heart failure (binary, yes [I50] or no). Laboratory biomarkers encompassed alanine transaminase (ALT; continuous, U/L), creatinine (continuous, umol/L), C-reactive protein (CRP; continuous, mg/L), glucose (continuous, mmol/L), glycated hemoglobin (HbA1c; continuous, mmol/mol), low-density lipoprotein cholesterol (LDL; continuous, mmol/L), and triglycerides (continuous, mmol/L).

### Outcomes

Outcome of this study was defined as a time-to-event variable, with follow-up commencing at the date of enrollment and terminating at the first occurrence of the outcome events or at the censoring date (i.e., December 1, 2024). In the PAH incidence cohort, the primary endpoint was the incidence of clinical diagnosis of IPAH; in the mortality cohort, the primary endpoint was defined as the all-cause mortality.

### Proteomic epidemiological analyses

Two steps cross-validated epidemiological analyses were performed to discovery robust plasma protein associated with outcomes. First, we constructed prospective cohort design and performed multivariable cox regression models within both PAH incidence and mortality cohort. Subsequently, for each cohort, case-control studies were designed using the 1:1 propensity score matching (PSM) (i.e., with vs. without diagnosed PAH in incidence cohort; deceased vs. survival in mortality cohort) and multivariable logistic regressions were applied. Each of the 2,920 proteins was modeled as an independent variable, adjusting for abovementioned covariates. To account for multiple testing in the high-dimensional discovery phase using cox regression models, proteins with a false discovery rate (FDR)-adjusted value < 0.05 were considered significant. In contrast, a nominal p-value < 0.05 was used as the significance threshold in the case-control validation due to the smaller number of comparisons and to preserve statistical power. Significant proteins both in cohort and case-control design with same directional effect were considered as robust proteins and subsequently analyzed.

Moreover, subgroup analyses were conducted in males and females using the same two-step methods, owing to the sex-dependent discrepancy in PAH onset and prognosis. The union of proteins identified in the overall population and in male and female subgroups was considered as the final set for each cohort.

### Enrichment analyses

To elucidate the biological pathways and functions of IPAH-associated proteins and the core pathway-participating proteins that define the dominant molecular subtypes, enrichment analyses were performed using the Gene Ontology (GO), Kyoto Encyclopedia of Genes and Genomes (KEGG), and Reactome databases. To ensure statistical rigor, these analyses were conducted against a given background reference set comprising the 2,920 proteins profiled in our study. Statistical significance was assessed via a hypergeometric test with FDR-adjusted *p*-value[<[0.05, and the top 10 GO terms, top 3 KEGG pathways, and top 10 Reactome pathways were reported.

### Proteomic cis-Mendelian randomization analysis

To explore the potential causal proteins associated with PAH, two-sample cis-MR analyses were conducted. Genetic instruments were constructed from cis-acting protein quantitative trait loci (cis-pQTLs) identified in 35,571 participants from the UKB-PPP. For each protein, the lead cis-pQTL, the variant most strongly associated within ± 500 kb of the protein-coding gene, was selected if it reached genome-wide significance (P < 5 × 10[[). To ensure instrument independence, variants in linkage disequilibrium were pruned at r² < 0.001 using European reference data from Phase 3 of the 1000 Genomes Project.

Summary statistics for outcome of diagnosed PAH were obtained from genome-wide association studies (GWAS) conducted in the FinnGen study ^43^. The cis-MR analyses employed five methods, with IVW or Wald ratio when only one SNP was available used for primary estimation, and Maximum Likelihood, MR-Egger, and Weighted Median applied as supplementary approaches. Each MR analysis utilized a single lead cis-pQTL per protein as the genetic instrument and causal estimates were considered statistically significant at a nominal p-value < 0.05. All analyses were restricted to individuals of European ancestry.

Sensitivity analyses for cis-MR results were conducted to account for directional horizontal pleiotropy and to obtain outlier-robust estimates. Directional horizontal pleiotropy was evaluated using the MR-Egger intercept for analyses with at least three instrumental variables, and results with a significant intercept (p-value < 0.05) were excluded to ensure the robustness of causal estimates. For outlier-robust estimation, proteins were required to show directionally consistent effects between IVW and other supplementary methods; otherwise, proteins failing this criterion were excluded. Additionally, we performed two-sample MR using cis-pQTLs from Fenland and deCODE as genetic instruments, with complementary datasets as outcome. Significant external proteins showing directionally consistent effects and overlapping with the significant MR results from UKB-derived proteins were defined as high-confidence proteins.

### Colocalization

To investigate shared genetic causality, we performed colocalization analyses between the cis-pQTLs of each protein and PAH. For each protein, we tested whether its cis-pQTLs shared a causal variant with PAH within a 500-kb genomic window. The analyses used default prior probabilities: p1 = 10[[ (prior probability of a genetic association for the protein), p2 = 10[[ (for PAH), and p12 = 10[[ (for both traits sharing a single causal variant). Proteins with posterior probability of a shared causal variant (PPshared) > 0.5 was considered to have evidence of colocalization.

### Clustering analysis

To identify molecular subtypes associated with high mortality risk in IPAH patients, we performed unsupervised clustering for clinical patients with IPAH at baseline in the mortality cohort and preclinical patients without IPAH at baseline in the incidence cohort using all cis-MR validated causal proteins. Before clustering, principal component analysis (PCA) was applied to reduce the dimensionality of these proteins, retaining the first two principal components for clustering. The optimal number of clusters was determined using the elbow and silhouette methods, which ultimately identified two clusters. (Supplementary Table.2) We selected the k-means algorithm, which iteratively assigns each participant to the nearest centroid and updates centroids as the mean of all points in each cluster until convergence, thereby minimizing the total within-cluster sum of squares, thereby grouping IPAH patients into two clusters. Moreover, Kaplan–Meier survival curves stratified by sex and proteomic cluster were plotted and compared using the log-rank test to evaluate cluster-based risk stratification for mortality. To visualize the proteomic signatures of molecular subtypes and their sex-specific patterns, we generated two sets of heatmaps. First, we constructed sex- and cluster-stratified heatmaps of mean z-scores (−1 to 1) for proteins associated with key biological pathways identified by KEGG and Reactome enrichment analyses, enabling the characterization of dominant pathway signatures for each sex and molecular subtype. Second, we created an individual-level heatmaps of truncated z-scores (−1 to 1) for all cis-MR–validated proteins, stratified by sex and molecular subtypes, to examine the relative expression patterns.

### Prediction models

To rank the significance and sex-dimorphic discrepancy of cis-Mendelian validated causal proteins, eXtreme Gradient Boosting (XGBoost)-based prediction models for PAH incidence and mortality were developed. To ensure robust performance estimation and prevent overfitting, we employed a nested cross-validation framework. This consisted of a 5-fold outer loop for unbiased performance evaluation and a 5-fold inner loop for feature selection (via LASSO regression) and hyperparameter tuning. We constructed clinical, proteomic, and combined risk models for both PAH incidence and mortality in the overall population as well as in male and female subgroups. Model discrimination was assessed using the area under the receiver operating characteristic curve (AUC), derived from the aggregated predictions of the outer validation folds, and compared using DeLong’s test. For biological interpretability, a final consensus model was retrained on the entire dataset using the optimal hyperparameters identified during cross-validation. SHapley Additive exPlanations (SHAP) values were then calculated across the full cohort to quantify the global contribution of each protein. The ranked importance of these proteins in the overall population and their sex-specific divergence was visualized using Sankey plots.

### Potential therapeutic targets

Robust proteins with causal effects on both PAH incidence and mortality were considered candidate therapeutic targets and classified into three tiers according to the strength of supporting evidence (i.e., targets supported by colocalization evidence, targets supported by multisource GWAS evidence, and targets supported by UKB-based GWAS evidence). Based on literature review, these candidates were further categorized into vascular-related and cardiac-related groups to facilitate downstream mechanistic investigation.

Given that functional interactions among proteins and related molecules may also modulate biological effects, the druggable target set was further expanded to include functionally connected receptors, ligands, or interacting proteins. To systematically identify approved drugs acting on these targets, three curated drug–target databases, DrugBank (https://www.drugbank.ca), Therapeutic Target Database (TTD, https://db.idrblab.net/ttd/), and Open Targets (https://platform.opentargets.org), were queried. All protein targets, interaction-based related targets, approved drugs, and therapeutic annotations were manually curated to support drug-repurposing analyses and future therapeutic development.

### Statistical analysis

All epidemiological analyses and data visualizations were performed in R (version 4.2.2). A two-sided FDR-adjusted p-value or nominal p-value was considered statistically significant according to the predefined analytical design. Missing NPX of proteins were imputed using a chained random forest algorithm with age and sex as predictors and Missing values in baseline covariates were imputed using the random forest-based multiple imputation implemented in the mice package. Multivariate cox proportional hazards models were applied for protein discovery, and multivariate logistic regression models were conducted after 1:1 PSM using the MatchIt package for epidemiological validation. Functional enrichment analyses were implemented using the clusterProfiler, org.Hs.eg.db, and ReactomePA packages. Two-sample cis-Mendelian randomization analyses were conducted using the TwoSampleMR package in R (version 4.1.3) to assess potential causal relationships between proteins and disease outcomes. For clustering analyses, PCA was first applied for dimensionality reduction, followed by an unsupervised k-means algorithm to identify distinct protein expression clusters. Kaplan–Meier survival curves and log-rank tests were used to compare the clinical outcomes across clusters. Prediction models for clinical outcomes were developed using XGBoost with LASSO-based feature selection, five-fold cross-validation, and model interpretation via SHAP values. Model performance was evaluated by the AUC and compared using DeLong’s test.

## Data Availability

All data produced are available online at UK biobank. (https://www.ukbiobank.ac.uk)

## Supplementary file

**Supplementary Figure.1.**
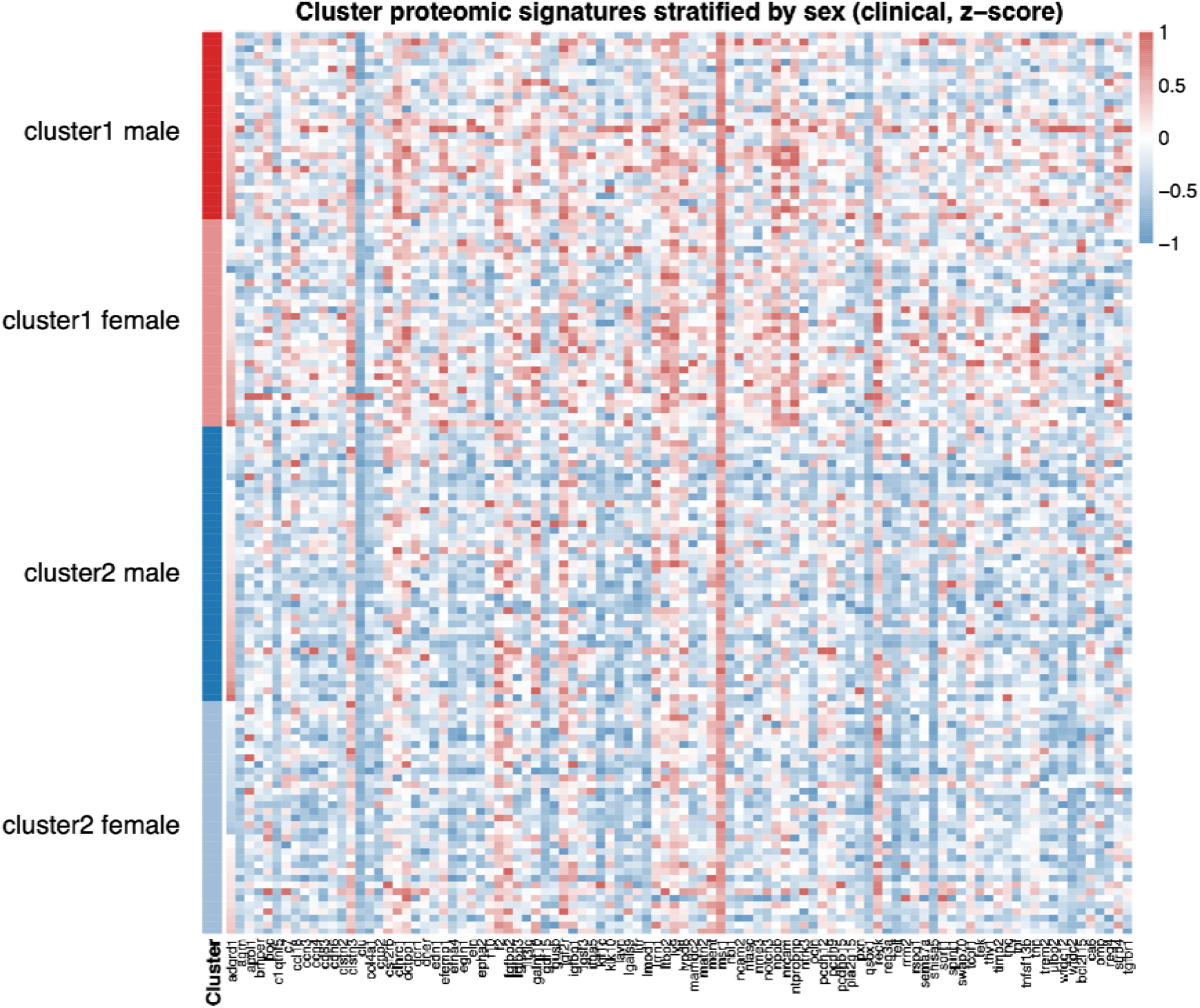
Sex-and cluster-stratified proteomic signatures for clinical patients with IPAH.

**Supplementary Figure.2.**
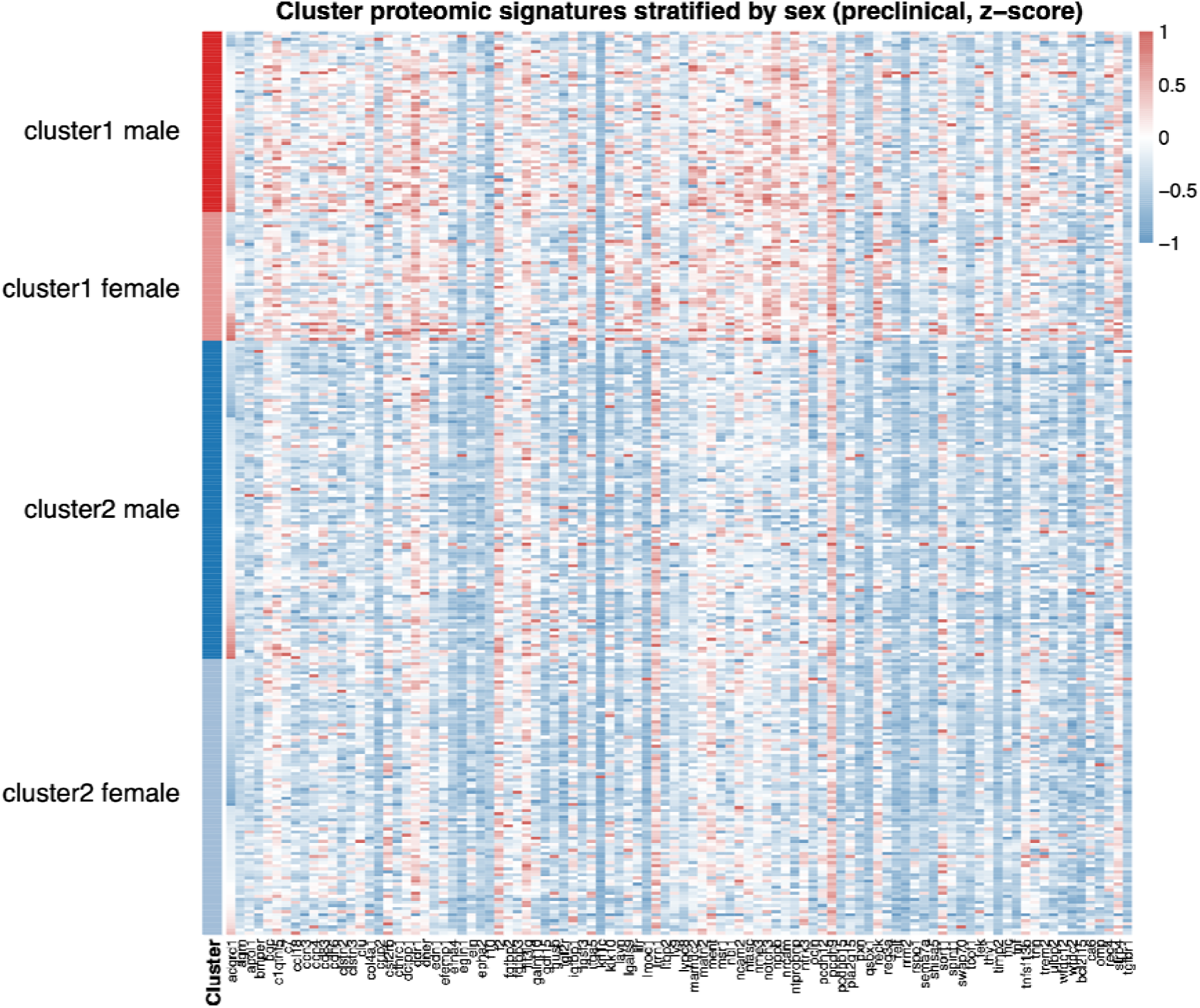
Sex-and cluster-stratified proteomic signatures for preclinical patients without IPAH.

**Supplementary Table 1.**
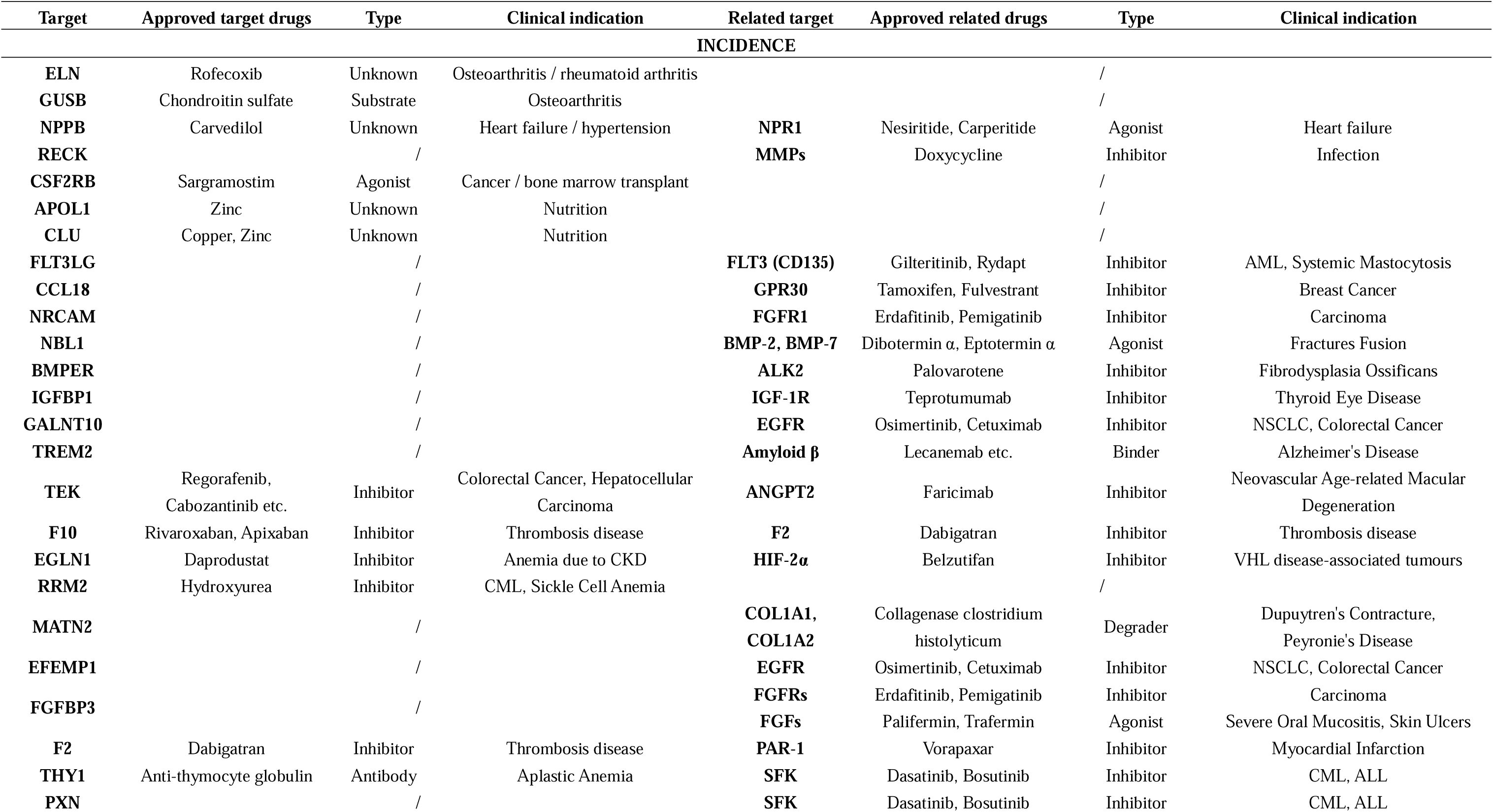
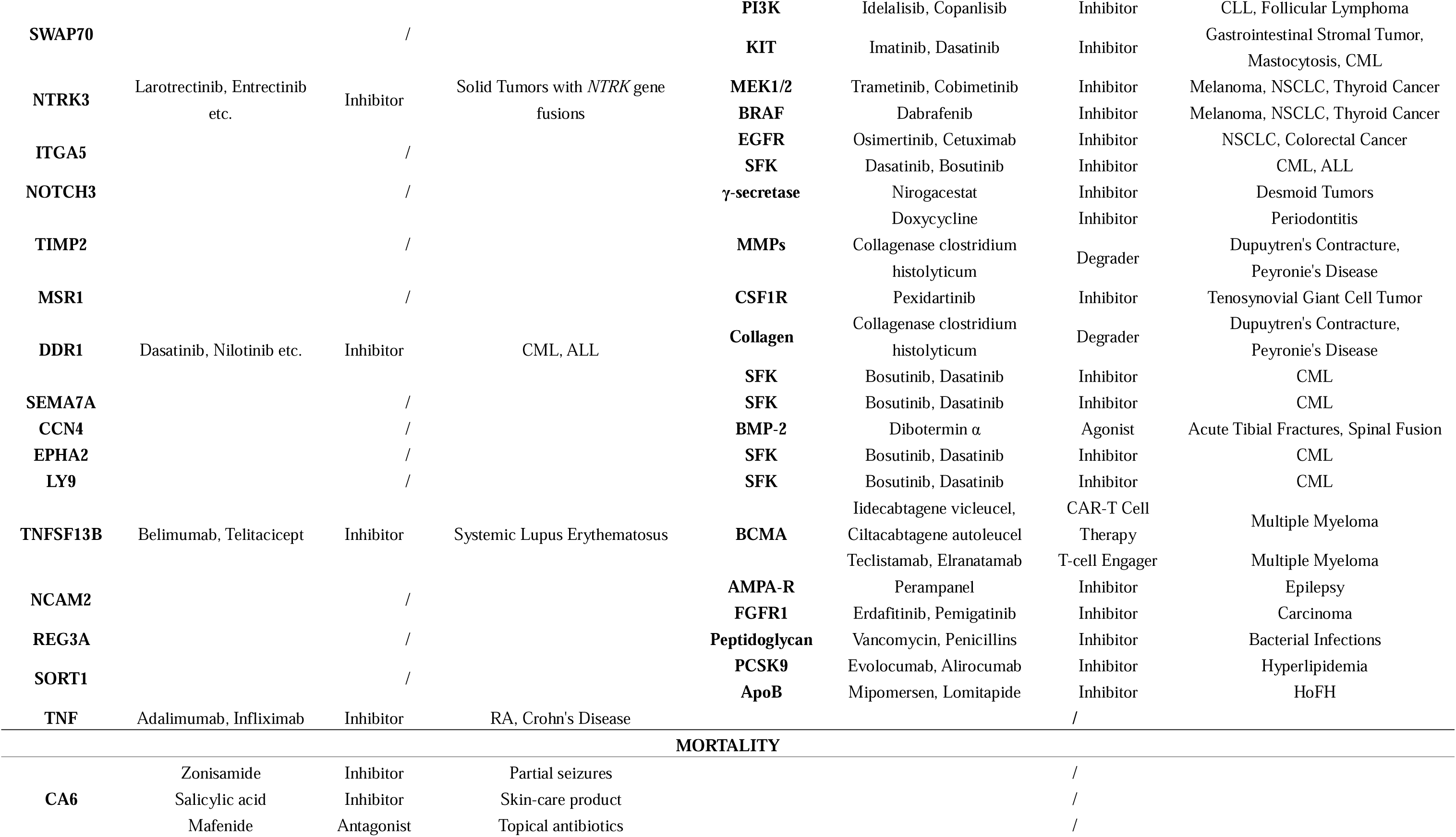
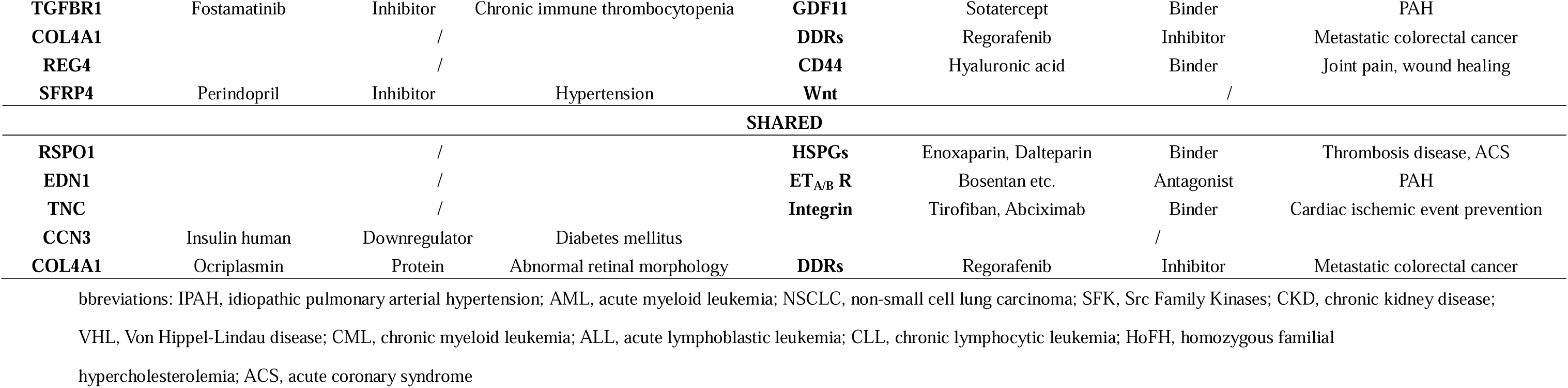
Target-drug and related target-drug annotation for the robust proteins associated with IPAH incidence and mortality.

**Supplementary Table 2.** Determination of cluster number.

| **Clinical patients with IPAH (mortality cohort)** |  |  |  |  |
| --- | --- | --- | --- | --- |
|  | k =1 | k = 2<sup>1</sup> | k = 3 | k = 4 |
| WSS | 4057.632 | 3262.522 | 3006.903 | 2845.785 |
| Silhouette value | / | 0.16671409 | 0.08615349 | 0.09447070 |
| **Preclinical patients without IPAH (incidence cohort)** |  |  |  |  |
|  | k =1 | k = 2<sup>1</sup> | k = 3 | k = 4 |
| WSS | 10513.832 | 8176.718 | 7606.408 | 7016.685 |
| Silhouette value | / | 0.20972652 | 0.10746615 | 0.12048164 |

<sup>1</sup>Number of clusters (i.e., k = 2 for clinical patients and k =2 for preclinical patients) were determined with the maximizing silhouette value and elbow method.
Abbreviations: IPAH, idiopathic pulmonary arterial hypertension; WSS, within-cluster sum of squares

